# Cine Cardiac MRI Captures Cardiovascular Disease Risk Beyond Established Clinical Risk Factors: Evidence from the UK Biobank

**DOI:** 10.64898/2026.08.10.26360086

**Authors:** Marta Hasny, Laura Daza, Keno Bressem, Maxime Di Folco, Julia A. Schnabel

## Abstract

Early and accurate risk stratification of cardiovascular disease (CVD) is crucial to initiate timely preventive interventions. As large-scale multimodal clinical cohorts become increasingly available, there is growing interest in whether incorporating additional sources of information can improve CVD risk stratification. Cine cardiac MR (CMR) represents a compelling example of such a source, as it captures objective, high-dimensional structural and functional information about the heart, independent of patient-reported data. In this study, we deploy a flexible vision-tabular method to incorporate cine CMR into CVD risk assessment together with structured clinical data. Using a large prospective imaging cohort from the UK Biobank, we show that cine CMR encodes CVD risk beyond established risk scores, increasing AUROC by 0.036 over SCORE2, the best-performing traditional risk score (0.742 vs. 0.706, *p* = 0.04). Furthermore, we find that cine CMR achieves risk discrimination capabilities on par with automated, image-derived phenotypes, removing the dependency on segmentation pipelines. Lastly, we demonstrate that integrating cine CMR with clinical variables through a vision-tabular learning framework stabilizes risk prediction under real-world conditions of incomplete tabular data, a common challenge in clinical practice. Together, these findings position cine CMR as a promising modality for CVD risk assessment.

## 1 Introduction

Cardiovascular diseases (CVD) remain the leading cause of death worldwide, accounting for millions of deaths each year [1, 2]. Effective CVD risk prediction is therefore critical for identifying high-risk individuals, enabling timely preventive interventions, and reducing disease burden. Established risk scores [3, 4], endorsed by clinical guidelines [5, 6], rely on a predefined set of structured attributes. While effective, these models are inherently constrained by their fixed input schema and cannot incorporate additional patient information beyond the preselected attributes. Meanwhile, a patient’s clinical information is not restricted to those few risk factors and can be extended to patient’s lifestyle information, medical or family history, or imaging data. Previous work on CVD risk assessment has shown that leveraging richer patient data, including broader health indicators and lifestyle factors, can improve risk estimation performance [7–9]. However, scaling risk assessment to larger, more heterogeneous attribute sets introduces a fundamental challenge: missing clinical information is an inherent characteristic of real-world healthcare data, and expecting hundreds of attributes to be consistently available for each subject undergoing risk assessment is unrealistic. Recent work has recognized this limitation, for example AdaCVD [10] demonstrated that LLM-based approaches can enable robust inference under variable numbers of attributes. All those advancements are however confined to extending the number of attributes they incorporate, but none extends to fundamentally different sources of information, such as medical imaging. Meanwhile, there has been growing interest in integrating imaging into CVD risk assessment as it can provide complementary information about the patient’s risk status [11]. Cine cardiac MRI (CMR) is a widely available, contrast-free imaging sequence that serves as the clinical reference modality for quantifying cardiac morphology and function [12], motivating studies evaluating its prognostic capabilities. Recent studies on the UK Biobank have shown that hand-engineered CMR radiomics features demonstrate incremental predictive value for incident CVD outcomes when added on top of vascular risk factors and conventional CMR indices [13]. Similarly, a different study showed that incorporating cardiac IDPs on top of non-image tabular data improves CVD risk stratification [14]. Driven by these findings, deep learning-based approaches have attempted to transition from manual feature engineering to automated, image-based risk assessment. For instance, CTSL [15] integrated cine CMR sequences with structured variables to perform survival analysis for major adverse cardiac events. Nevertheless, a critical gap limits the translation of these methodologies into standard clinical practice. Current multimodal deep learning frameworks, and comparative conclusions drawn by such works, are generally evaluated exclusively in idealized settings where all data modalities are perfectly complete. They fail to account for the attribute missingness and upstream pipeline failures inherent to real-world healthcare environments. Furthermore, they do not compare against established clinical risk scores such as Framingham [3] and SCORE2 [4] or their underlying risk factor attributes, limiting the ability to contextualize the added prognostic value of imaging relative to the clinical information already used in standard practice.

Meanwhile, recent advances in vision-tabular learning have enabled architectures that handle heterogeneous and missing inputs natively at inference [16–18]. Some of those advancements have been based on tabular foundation models [19–21], which support missing values, and compared to aforementioned LLMs, account for millions of parameters instead of billions. This multimodal integrations opens up the possibility of risk assessment frameworks that flexibly incorporate rich clinical data, including cardiac imaging, without imposing strict constraints on data availability at prediction time. Furthermore, several of these methods have been specifically developed for and evaluated on cine CMR workflows [16, 18], demonstrating significant promise for cardiac applications and providing robust pretraining strategies for visual encoders that adapt well to downstream tasks, even on limited datasets. Nevertheless, the tasks analyzed in literature so far have remained strictly limited to static cardiac diagnoses using cine CMR alongside tabular data. These multimodal models, however, present broader clinical opportunities: they can actively support long-term clinical decision-making by enabling flexible risk assessment, while simultaneously serving as a powerful tool for investigating the predictive contributions of imaging and structured clinical data, both individually and jointly.

In this study, we advance vision-tabular learning to the task of CVD risk assessment, developing a multimodal classification framework supporting image-only, tabular-only, and joint multimodal inference under missing values. We leverage this framework to systematically evaluate the prognostic capabilities of cine CMR, both directly as images and through image-derived phenotypes (IDPs). Our findings demonstrate that cine CMR enhances cardiovascular risk prediction beyond established risk scores. Furthermore, we find that the images can substitute for automated IDPs, capturing equivalent prognostic information and thus removing the dependency on segmentation pipelines, with interpretability analyses confirming that the model attends to clinically meaningful cardiac structures. Lastly, we find that in the vision-tabular setting with hundreds of clinical attributes, imaging improves model robustness to missing or erroneous values. This finding suggests that cine CMR can provide a reliable anchor for risk assessment in real-world clinical settings where complete and error-free patient records cannot be guaranteed.

## 2 Results

### 2.1 CVD Risk Assessment in UK Biobank

We investigated the prognostic value of imaging data for CVD risk assessment by conducting a study using participants from the UK Biobank [23] imaging cohort (first assessment only). We first defined the tabular feature set used in our analysis. In line with established clinical risk scores [3, 4], we constructed a set of basic CVD risk factors comprising age, sex, total cholesterol, high-density lipoprotein (HDL) cholesterol, blood pressure (BP) medication use, systolic BP, smoking status, and diabetes. We further extracted IDPs, such ejection fraction, cardiac index, or cardiac output, from cine CMR images using a validated segmentation pipeline [22]. These variables, together with additional clinical attributes collected in the UK Biobank cohort, define the list of features used in this study. Specifically, we considered eight categories: (1) basic CVD risk factors, (2) ECG conduction metrics, (3) pulse wave analysis (PWA) parameters, (4) medical history, (5) lifestyle factors, (6) physical measurements, (7) family history, and (8) IDPs. A complete list of non-CMR attributes is provided in Appendix Tabel A2, while the IDPs are listed in the Appendix Tabel A3.

The selected cohort comprised 49,737 participants with short-axis cine CMR examinations acquired between 2014 and 2022. Participants with a CVD diagnosis prior to or within one year of the imaging date, as well as those with less than one year of available follow-up, were excluded. Consistent with prior UK Biobank cardiovascular risk prediction studies [7, 10], we defined a composite CVD outcome using ICD-10 diagnoses of ischemic heart disease (I20-I25), heart failure (I50), cerebrovascular disease (I60-I69), and vascular dementia (F01). Participants who developed one or more of these outcomes during follow-up were classified as at-risk (Fig. 1b). Incident events were evaluated over the maximum available follow-up period. Since imaging data were acquired between 2014 and 2022 and diagnosis records in our cohort were extracted in 2022, event-free participants had variable follow-up durations of up to 8 years. Participants without a recorded CVD diagnosis by the extraction date were treated as event-free and incorporated through right-censoring. The final cohort was partitioned into training (N=1,428, balanced 50% at-risk), validation (N=2,366), and test (N=5,961) sets (Table 1).

**Fig. 1.**
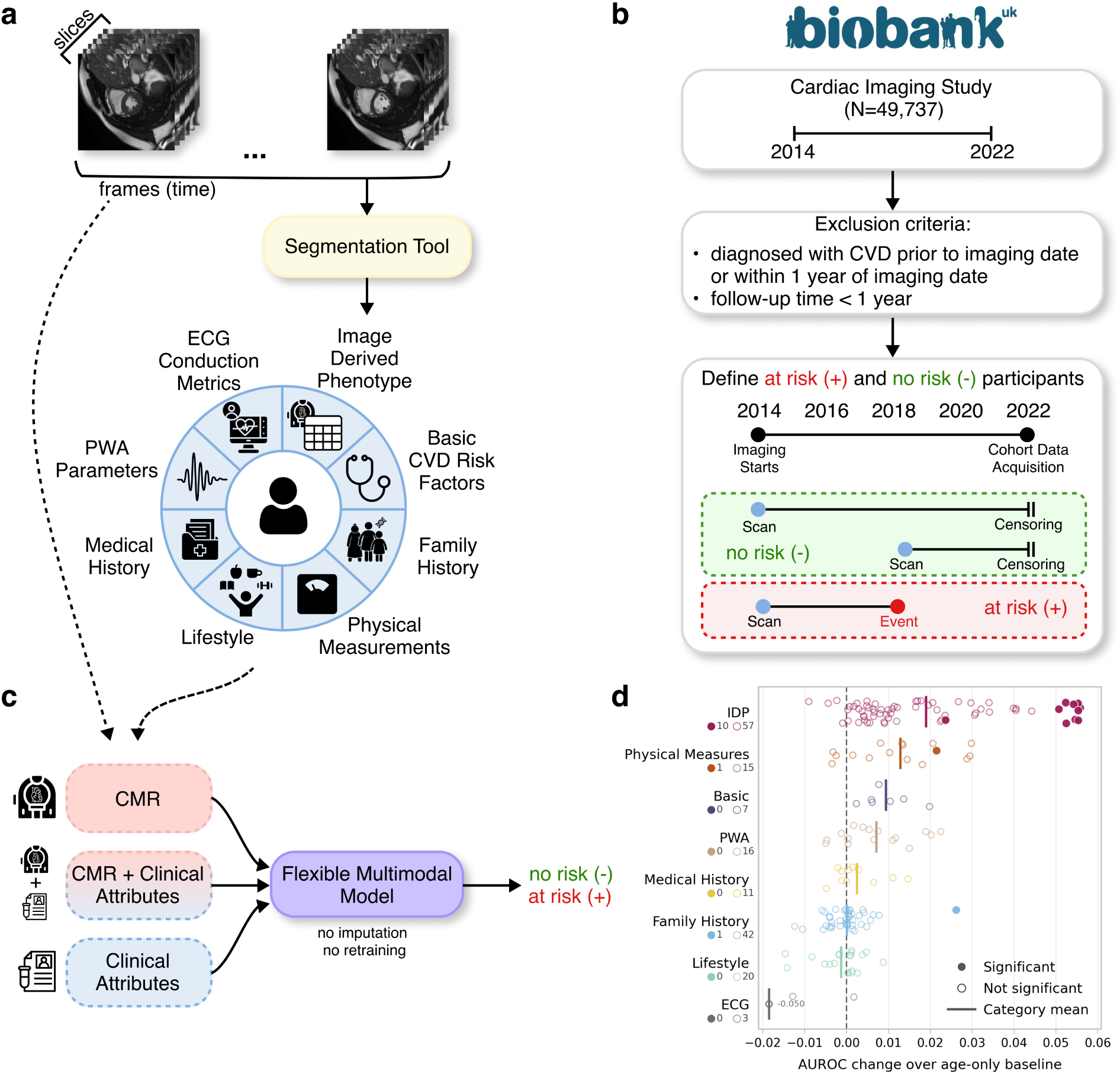
Study design and data overview. (a) Cine CMR acquisition and clinical attribute categories used in this study. IDPs are extracted from cine CMR using an automated segmentation tool [22]. (b) UK Biobank cohort selection and outcome definition. At-risk (+) participants developed a CVD outcome post-imaging; no risk (−) participants have no recorded CVD event within available follow-up time. (c) The flexible multimodal vision-tabular pipeline accepts CMR imaging, clinical attributes, or both without retraining or imputation. (d) Individual attribute predictive capability against age-only baseline (AUROC 0.654*±* 0.023). For each candidate attribute, a classifier (TabPFN 2.5 [20]) was trained on age plus given attribute and evaluated against an age-only baseline. Points show the AUROC change relative to the age-only baseline for each attribute, grouped and color-coded by attribute category. We provide the names of the attributes that achieved significant results in the Appendix Tabel A1.

**Table 1.** Cohort characteristics stratified by CVD risk status across dataset splits. Demographic and clinical characteristics of participants in the training, validation, and test set, stratified by CVD risk status (at-risk vs. no risk). Follow-up time reflects time from imaging to first CVD event for at-risk participants and time from imaging to censoring for healthy participants. BP: blood pressure.

|  | Train (N=1,428) |  | Validation (N=2,366) |  | Test (N=5,961) |  |
| --- | --- | --- | --- | --- | --- | --- |
|  | At-Risk | No risk | At-Risk | No risk | At-Risk | No risk |
| Number of Subjects | 714 | 714 | 71 | 2,295 | 140 | 5,821 |
| Women [%] | 37.96 | 54.76 | 32.40 | 52.64 | 36.43 | 54.77 |
| Body Mass Index (BMI) | 26.84±4.10 | 26.48±4.32 | 27.80±5.37 | 26.38±4.33 | 27.26±4.31 | 26.31±4.22 |
| Age [Years] | 67.09±7.25 | 64.43±7.72 | 68.32±7.32 | 64.14±7.74 | 68.19±6.35 | 64.09±7.63 |
| Smoking [%] | 5.46 | 2.1 | 2.82 | 3.49 | 3.57 | 3.3 |
| Systolic BP | 145.26±19.26 | 140.53±19.54 | 145.77±21.82 | 140.05±19.42 | 149.01±21.61 | 139.44±19.37 |
| BP Medication [%] | 28.43 | 21.01 | 26.76 | 19.96 | 35.0 | 19.91 |
| Total Cholesterol [mmol/l] | 5.77±1.09 | 5.80±1.03 | 5.72±1.07 | 5.77±1.07 | 5.70±1.07 | 5.73±1.04 |
| Diabetes [%] | 4.48 | 1.82 | 7.04 | 2.79 | 4.29 | 2.44 |
| Follow-up Time [Years] | 3.39±1.83 | 4.13±1.41 | 2.62±1.80 | 4.46±1.52 | 2.69±1.85 | 4.44±1.51 |

To evaluate the prognostic value of cardiac imaging across variable clinical settings, we advanced a flexible vision–tabular deep learning framework [18], extending its ability to natively handle image-only and joint multimodal inference to also support a tabular-only setting. For imaging input, we constructed cine CMR volumes consisting of 10 temporal frames across 11 short-axis slices per subject, which are provided directly to the model. All inference schemes are performed without requiring missing-data imputation or model retraining. We do not perform imputation at training. An initial analysis (Fig. 1d) of the independent predictive performance of each individual clinical and imaging attribute revealed that IDPs possess strong predictive capability for CVD risk. This provides clear evidence that cardiac imaging carries prognostic value, which directly motivated our subsequent evaluation of how to best leverage cine CMR sequences for robust, real-world cardiovascular risk assessment.

### 2.2 CVD Risk Prediction Performance

We first evaluate model performance in a controlled setting in which each tabular model is trained on the specific set of clinical attributes it is evaluated on. We refer to these as *specialized* models throughout the remainder of the paper. In the vision–tabular setting, the multimodal model is trained once using all available attributes with TabPFN [20]. For evaluation, the tabular encoder is then replaced at inference time with the corresponding specialized model without any retraining. We perform the evaluations on three tabular attribute sets: the *basic* set consists of established CVD risk factors as defined in the previous section, the *non-CMR* set includes all attribute groups shown in Fig. 1a except IDPs, and the *all* set includes all attributes used in this study, including IDPs.

#### 2.2.1 Performance Using Basic Risk Factors

Our multimodal model, incorporating basic clinical attributes and cine CMR, achieves the best performance across all tabular attribute settings (Fig. 2a), reaching an AUROC of 0.742. This significantly outperforms established clinical risk scores, including Framingham [3] (AUROC: 0.685, *p*=0.0003) and SCORE2 [4] (AUROC: 0.706, *p*=0.04). Because follow-up times in our cohort are variable and do not directly correspond to the fixed 10-year prediction horizon of Framingham and SCORE2, we additionally compare against a Cox proportional hazards model [24], which serves as the strongest tabular-only baseline and achieves an AUROC of 0.707. Our multimodal model again shows a significant improvement over this baseline (*p*=0.03). Finally, the multimodal model outperforms its tabular-only counterpart (AUROC: 0.707, p=0.02), where the same architecture is used without access to cine CMR. This demonstrates that cine CMR provides strong predictive capability for CVD risk assessment beyond what is captured by established clinical risk factors.

**Fig. 2.**
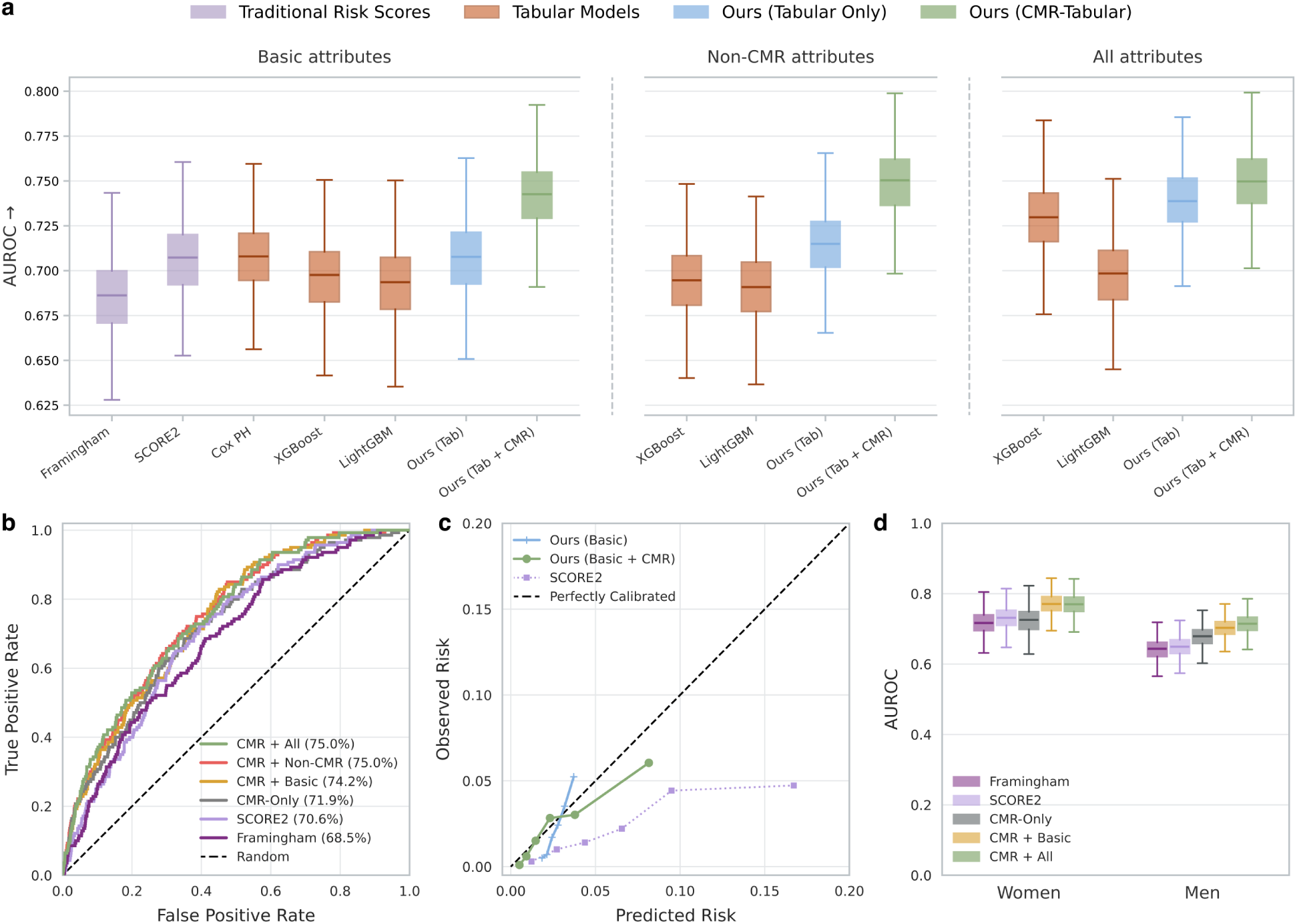
CVD risk prediction performance. (a) Bootstrapped AUROC distributions across attribute configurations for traditional risk scores, tabular baselines, and our method. We only use specialized tabular models for this evaluation, meaning they are always trained on the same attribute set that they are evaluated on. Our multimodal model was trained once and the tabular model is swapped for the specialized one without any retraining of the multimodal module. Non-CMR attributes contain all the attribute groups except IDPs, while all attributes incorporate IDPs on top of the no-CMR fields. (b) ROC curves for our vision-tabular model across input configurations. (c) Calibration curves evaluating the performance of our vision-tabular method for tabular-only input and CMR-tabular input against SCORE2. Extended figure incorporating Framingham is available in the Appendix Fig. A1. (d) Comparison of our vision-tabular framework against SCORE2 and Framingham stratified by sex.

#### 2.2.2 Incorporating Broader Clinical Attributes

Prior work has shown that incorporating a broader range of clinical attributes improves CVD risk prediction performance [7–9]. Consistent with this, we observe that performance increases as additional attribute groups are included across all evaluated methods (Fig. 2a), a pattern also reflected in the corresponding ROC curves (Fig. 2b). Notably, when using non-CMR attributes alongside either cine CMR images or IDPs as alternative representations of the imaging information, the vision-tabular model combining images with non-CMR attributes achieves a higher mean AUROC than the corresponding tabular-only model using non-CMR attributes and IDPs (AUROC: 0.750 vs. 0.739), although the difference is not statistically significant (*p* = 0.41). This suggests that cine CMR used directly provides prognostic information comparable to that captured by IDPs, without requiring an intermediate segmentation step. Furthermore, when CMR imaging is already included, adding IDPs on top of the non-CMR attributes does not further improve performance (AUROC: 0.75 for CMR + Non-CMR vs. 0.75 for CMR + All), indicating redundancy between IDPs and representations learned directly from cine CMR imaging. A direct comparison of IDP-based and cine CMR imaging-based performance across imaging-only and multimodal settings is provided in Appendix Fig. A2, further supporting our finding that using cine CMR imaging directly consistently matches or exceeds IDP-based predictions across all attribute configurations.

To assess whether improvements in discrimination translate into well-calibrated risk estimates, we evaluate model calibration (Fig. 2c). We use the basic attributes as the tabular input for our vision-tabular risk estimator to ensure direct comparison against SCORE2. For visual clarity, Framingham is excluded from the main figure and the extended set of calibration curves is provided in Appendix Fig. A1. Both the tabular-only and the vision-tabular versions of our method showed improved calibration over SCORE2, that tends to overestimate the risk across the predicted range. The basic + CMR model closely tracks the diagonal of perfect calibration across low-to-moderate predicted risk, outperforming the tabular-only model, which diverges from the diagonal earlier by overestimating risk in this range. At the highest predicted risk values, the basic + CMR model shifts toward overestimation, similar in direction to SCORE2 though considerably smaller in magnitude, while the tabular-only model instead underestimates risk in this range.

We further stratify performance by sex (Fig. 2d). All models achieve higher AUROC in women than in men, a pattern consistent across both traditional risk scores and CMR-based models. Importantly, the CMR + basic configuration consistently outperforms the traditional risk scores, Framingham and SCORE2, indicating that the discriminative advantage of incorporating cine CMR imaging is not sex-specific. Extended subgroup analyses across diabetes, age, BMI, and smoking status are provided in Appendix Fig. A3, and consistent with the pattern observed here: CMR-based models consistently match or outperform traditional risk scores across subgroups.

### 2.3 Robustness Analysis

#### 2.3.1 Adapting to Missing Values

In real-world clinical settings, large-scale attribute sets are often incomplete, and models must therefore remain robust to missing input data. We first evaluate the robustness of our vision-tabular pipeline to attribute missingness by simulating fixed subset of features being consistently available at inference (basic attributes, non-CMR attributes, and all attributes). We compare models trained on all available attributes with those trained and evaluated on the same subset (*specialized* models; Fig. 3a). In this setting, we assess whether the vision-tabular framework maintains performance when specific attribute groups are removed. We find that incorporating cine CMR stabilizes model performance under structured attribute missingness. In the tabular-only configuration, removing IDPs leads to a substantial performance drop, even though all other clinical attribute groups remain available (AUROC: 0.739 vs. 0.687, p=0.0004). This is notable given that IDPs account for 7 of the 20 most influential predictors based on SHAP analysis (Fig. 4a). In contrast, in the multimodal configuration, performance remains stable when IDPs are removed (AUROC: 0.75 vs. 0.742, p=0.06). Importantly, across these structured missingness conditions, the multimodal model does not fall below the performance of established clinical risk scores, including Framingham and SCORE2, as opposed to its tabular-only counterpart. Calibration results under structured missingness, assessed via the Brier score, are provided in the Appendix Fig. A4.

**Fig. 3.**
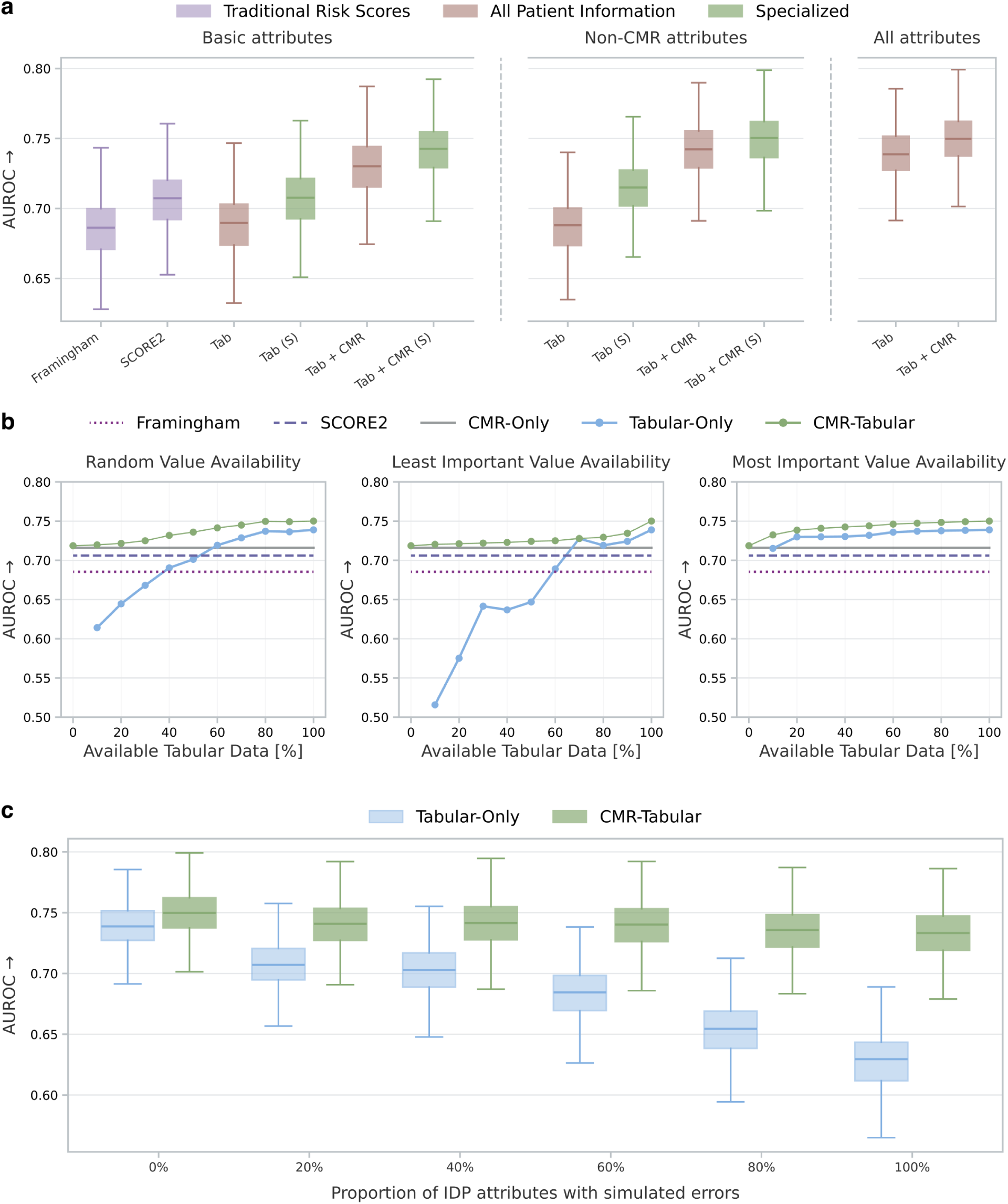
Robustness analysis. (a) Performance of specialized versus non-specialized (trained on all the attributes) models across attribute configurations. (b) Performance across different tabular data availability percentages over three missingness scenarios: random, least important, and most important. SCORE2, Framingham, CMR-only model (ResNet-50 [25]) are used as reference lines. (c) Robustness to simulated errors in IDP attributes at increasing proportions of corrupted values, with and without CMR used as a direct input.

**Fig. 4.**
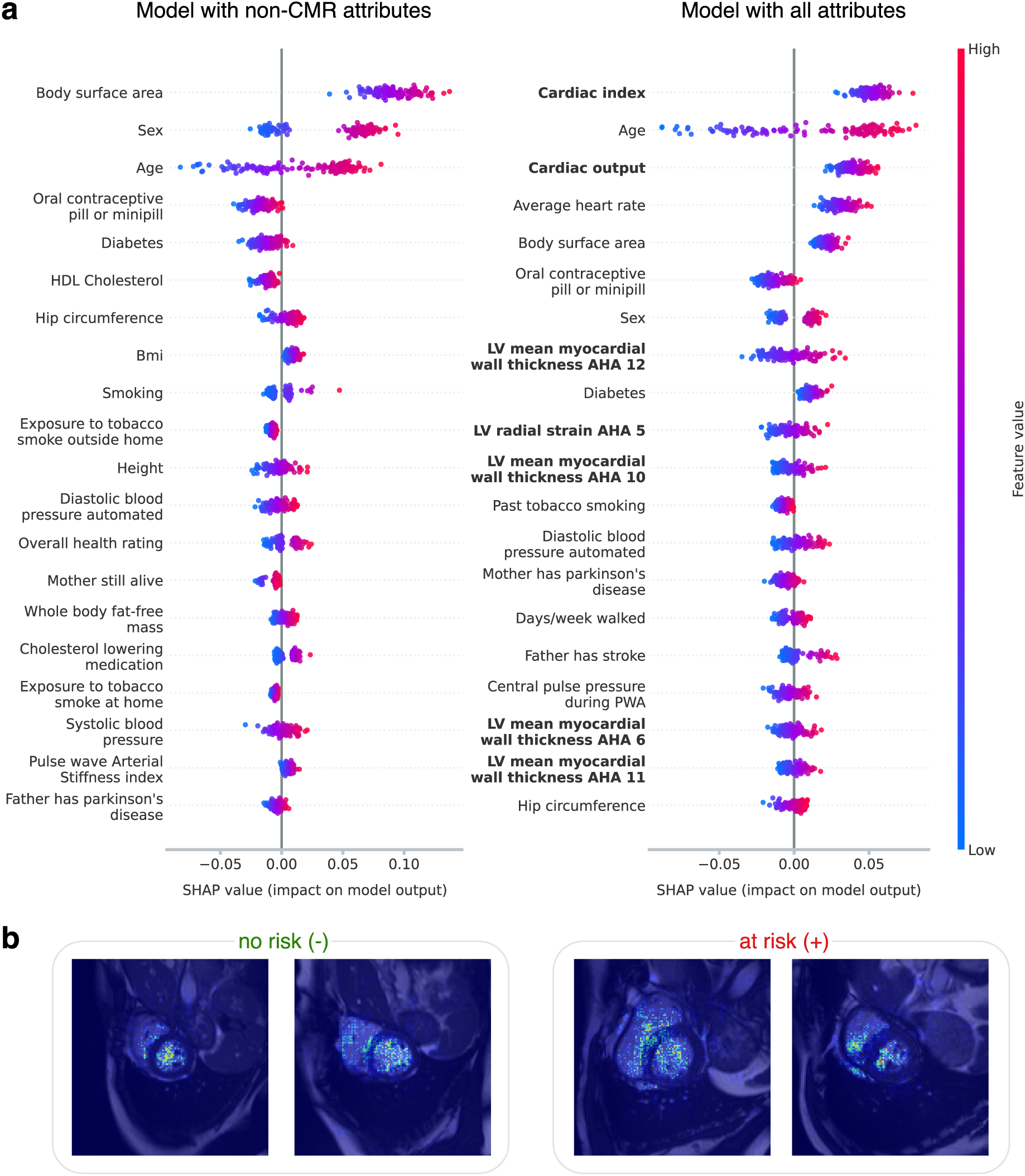
Interpretability analysis. (a) SHAP feature importance for models trained using non-CMR attributes (left) and all attributes (non-CMR + IDPs; right). CMR-derived attributes (bold) dominate when included. (b) FullGrad [28] saliency maps for representative no risk and at-risk participants. Images reproduced by kind permission of UK Biobank ©.

We further evaluate robustness to missing tabular data by varying attribute availability from 0–100% (Fig. 3b). We consider three missingness scenarios: (1) random attribute availability (sample-level missingness), (2) availability of the least important attributes, and (3) availability of the most important attributes (both column-level missingness). Attribute importance was determined using SHAP analysis [26] (Fig. 4a). We compare the vision-tabular and tabular-only configurations against a CMR-only baseline (3D ResNet-50 [25]), SCORE2 [4], and Framingham [3]. At 0% tabular availability, the CMR-tabular configuration reduces to the CMR-only setting. The tabular-only configuration struggles to match the performance of SCORE2, the best performing traditional risk score, under random and least-important-attribute availability, requiring approximately 60% of attributes to be present before reaching comparable performance. Robust performance is only observed when the most important attributes are preferentially retained, highlighting the strong dependence of the tabular model on a relatively small subset of highly informative features. In contrast, the vision-tabular configuration consistently outperforms Framingham, SCORE2, and the CMR-only baseline across all levels of attribute availability and under all missingness scenarios. These results demonstrate that cine CMR imaging provides a stabilizing predictive signal when clinical tabular information is incomplete.

#### 2.3.2 Performance under Attribute Corruption

Prior work has shown that the performance of cardiac segmentation pipelines can vary across demographic groups [27]. Errors in automated segmentation pipelines can introduce downstream noise into IDPs, potentially degrading the performance of models that rely on them. To evaluate whether cine CMR can mitigate the impact of such errors, we progressively corrupted ground-truth IDP values by shifting a randomly selected, nested subset of attributes (20-100%) by 3-4 standard deviations from the population mean per subject, simulating strong errors while keeping corrupted values within physiologically plausible ranges (Fig. 3c). Across increasing levels of attribute corruption, the performance of the vision-tabular configuration remained largely stable despite minor fluctuations (ΔAUROC = −0.017). In contrast, the tabular-only configuration exhibited a substantially larger degradation (ΔAUROC = −0.111). While corruption of IDPs directly impacts the tabular input representation, the multimodal configuration can continue to leverage information directly from the image, preserving performance when segmentation-derived measurements become unreliable, even under strong errors.

### 2.4 Interpretability Analysis

Understanding the drivers of model predictions is essential for clinical applications of machine learning. As our approach integrates both visual and tabular inputs, we perform separate interpretability analyses for each modality.

#### 2.4.1 Tabular Interpretability Analysis

One of the motivations of our study was the finding that IDPs exhibit strong predictive performance for CVD risk assessment (Fig. 1d). We therefore assess attribute importance under two configurations: with (all attributes) and without inclusion of IDPs (non-CMR attributes only), using SHAP analysis [26] (Fig. 4a). When IDPs are included, they account for 7 of the 20 most important attributes, further supporting their prognostic relevance. Notably, cardiac index and cardiac output emerge as the most influential predictors overall, surpassing age and sex, which are the dominant features in the non-IDP configuration and are well-established clinical risk factors. Several features that rank highly in the absence of IDPs, such as smoking, HDL cholesterol, and BMI, are displaced from the top-ranked attributes once IDPs are included, suggesting that part of their prognostic value could be captured by cardiac structural and functional measures. Age remains among the most important predictors in both settings, consistent with its established role in CVD risk stratification.

#### 2.4.2 CMR Interpretability Analysis

To examine which regions of the cine CMR contribute to model predictions, we generate saliency maps using FullGrad [28] for representative no risk and at-risk participants (Fig. 4b). Across both groups, the highest activations are consistently localized to cardiac structures, including the left and right ventricles, indicating that the model focuses on anatomically relevant regions rather than background areas. These findings suggest that the strong predictive performance of cine CMR observed throughout our experiments is likely driven by the model focusing on anatomically and physiologically meaningful cardiac structures, rather than spurious correlations or image noise.

## 3 Discussion

Cine CMR is the clinical gold standard for assessing cardiac function, yet its prognostic value for long-term CVD risk in individuals without prior cardiac events remains largely unexplored. In this study, we demonstrate that cine CMR encodes prognostic information beyond established risk factors for cardiovascular risk assessment. We advance vision-tabular multimodal learning [18], developing a risk classifier capable of inference from tabular data only, cine CMR only, or both modalities jointly. This design enables strong multimodal fusion while allowing us to evaluate the standalone predictive contribution of each modality as well as their combined value for cardiovascular risk assessment.

The number of CMR examinations has grown nearly fourfold over the past decade [29]. The growing number of existing scans creates a prospect of further leveraging those scans in an opportunistic manner. Our results support this possibility, showing that cine CMR captures CVD risk beyond established clinical risk scores. For patients already undergoing CMR, this prognostic signal can be extracted without additional acquisition cost or patient burden. Meanwhile, prior works on AI-driven CVD risk prediction has mainly focused on incorporating additional clinical attributes, such as lifestyle, family history, or genetics, as the input to the model [7–10]. While effective, this strategy requires hundreds of attributes spanning multiple clinical modalities, creating practical issues: the more attributes a model relies on, the more vulnerable it becomes to missing attributes inherent to real-world clinical setting. Our vision-tabular risk estimator was designed to operate at a similar scale of clinical attributes and our findings show that cine CMR improves robustness under incomplete clinical input. In contrast to tabular-only models, which fall below the performance of established risk scores under attribute missingness, the CMR-tabular configuration maintains performance above clinical baselines across all levels of attribute availability, including the complete absence of tabular data.

Cine CMR is an established sequence with a rich set of validated image-derived measures, such as ejection fraction, myocardial wall thickness, ventricular volumes, and cardiac mass [30]. Deriving these features reduces the high-dimensional cine CMR data into a compact set of well-established attributes that can be readily integrated with other clinical variables into a single tabular model. However, extracting IDPs requires manual delineation of the cardiac anatomy or automated segmentation pipelines, whose performance has been shown to vary across demographic groups [27], potentially introducing downstream bias. Meanwhile, recent works have proposed cardiac foundation models capable of processing high-dimensional cine CMR data directly [31–33]. Building on such pretrained model [18] as an image encoder, our results indicate that cine CMR can be used as a direct input without loss of performance, reducing dependence on upstream segmentation pipelines that could be prone to errors and degrade downstream predictions. Furthermore, interpretability analyses indicate that our model focuses on clinically relevant cardiac structures, suggesting that predictions are driven by meaningful anatomical signals rather than spurious correlations.

Despite these promising findings, several limitations should be acknowledged. Our study is retrospective and observational in nature. Diagnosis information in the UK Biobank is largely self-reported, meaning some participants may be unaware of an existing or developing condition, both at the time of imaging and during the follow-up period, and could therefore be misclassified as no risk subjects. As a result, it cannot be excluded that the model identifies earlier diagnosis of existing or subclinical disease rather than risk preceding disease onset. All analyses were conducted on the UK Biobank imaging cohort, which is predominantly white British and drawn from a volunteer population, limiting generalizability to more diverse clinical settings. External validation on independent cohorts would strengthen the conclusions, but remains constrained by the limited availability of large-scale datasets combining cine CMR with longitudinal CVD outcomes. In addition, follow-up durations vary and are shorter than the 10-year horizons used in clinical risk scores such as Framingham and SCORE2, making direct comparisons approximate.

In summary, our findings demonstrate that cine CMR, increasingly acquired for diagnostic cardiac assessment, encodes prognostic information that extends beyond established clinical risk factors and can be leveraged for cardiovascular risk stratification in an opportunistic manner. By advancing vision-tabular learning to this task, we show that end-to-end learning from cine CMR images matches the predictive performance of expert-derived imaging phenotypes without requiring upstream segmentation, and that the inclusion of cine CMR stabilizes risk prediction when clinical attributes are missing, inherent to real-world healthcare settings. Together, these results position cine CMR as a promising modality for cardiovascular risk assessment, and motivate prospective validation in clinically referred cohorts as a natural next step toward integration into routine cardiac imaging workflows.

## 4 Methods

### 4.1 UK Biobank

UK Biobank [23] is a longitudinal population study comprised of a cohort of 500,000 men and women from the UK aged 40-69 year at recruitment (2006-2010). The baseline assessment included comprehensive questionnaires, interviews, physical measures, and blood sampling, while the disease outcomes and mortality are continuously reported, tracked via electronic hospital records, cancer registries, death registries, and primary care records. Additionally, the UK Biobank imaging study was launched in 2014 with the goal of scanning 100,000 participants (20% of the original cohort). One of the modalities included in the imaging assessment is cine CMR [34, 35]. The cine CMR scans consist of a multi-view multi-slice 2D + time images in short-axis and long axis views. In this study we use the short-axis images, which typically consist of around 8-12 slices and 50 time frames per cardiac cycle.

Data collection by the UK Biobank was ethically approved by the North West Multi-centre Research Ethics Committee. As our study strictly utilizes this pre-existing data for secondary analysis, no additional ethical clearance is required. All participants provided written informed consent, and all records were fully de-identified prior to analysis. Qualified researchers can apply to access the data via the UK Biobank portal (www.ukbiobank.ac.uk). This research has been conducted using the UK Biobank Resource under Application Number 87065.

### 4.2 CVD Risk Dataset Construction

Our UK Biobank cohort consists of 49,737 participants with cardiac imaging data. The dataset was initially split into a training set of 39,975 participants, a validation set of 2,794 participants, and a test set of 6,968 participants. These initial splits were used for pretraining of the vision and tabular encoders [18], and included all participants regardless of CVD status. Following pretraining, participants diagnosed with CVD prior to imaging date were excluded from all splits. We define CVD as an occurrence of ischemic heart disease (ICD-10: I20–I25), heart failure (I50), cerebrovascular disease (I60–I69), or vascular dementia (F01), following the definition from prior works [7, 10]. A participant is labeled as at-risk if they develop a CVD diagnosis at least one year after the imaging appointment within the available follow-up period, and as no risk otherwise. This one-year exclusion window was applied to reduce the likelihood of including participants who may have had subclinical disease present at the time of imaging but were yet undiagnosed. Given that the imaging study was performed between 2014 and 2022 and the diagnosis data were downloaded in 2022, the maximum observed time between imaging and a recorded CVD event in our cohort is 8 years. To ensure sufficient observation time for reliable classification, participants with less than one year of follow-up between imaging date and diagnosis data download were excluded.

For each participant, we extracted 183 tabular attributes spanning eight categories: basic CVD risk factors, ECG conduction metrics, PWA parameters, medical history, lifestyle factors, physical measurements, family history, and IDPs. A complete list of attributes is provided in Appendix Tabel A2. For the imaging modality, we used short-axis cine CMR. To obtain a fixed-size input for the vision encoder, the temporal dimension was uniformly subsampled by retaining every fifth frame, resulting in 10 frames per cardiac cycle, while the slice dimension was padded or cropped to 11 slices. Each frame was subsequently cropped to a spatial resolution of 128 × 128 pixels centered on the left ventricle, yielding input volumes of shape 11*×*10*×*128*×*128.

As participants with CVD risk accounted for only 2.4% of the full training set, we balanced the training set to include all 714 CVD risk-positive samples alongside 714 randomly sampled healthy participants. The final validation set consists of 2,366 participants and the test set of 5,961 participants, both reflecting the natural class distribution of the cohort (2.5% at-risk).

### 4.3 Model Design and Training

We build our CVD risk prediction model on the state-of-the-art multimodal vision-tabular learning method, RoVTL [18]. We find vision-tabular models to be a suitable paradigm for CVD risk prediction involving imaging, as current risk factors can be represented in the form of tabular data. Furthermore, vision-tabular models have shown robustness to missing attributes at inference [16, 18]. However, previous works on vision-tabular learning have not accounted for the case when the imaging modality is missing. We adapt our vision-tabular method to account for this case by leveraging a tabular foundation model based on in-context learning, TabPFN 2.5 [20], as our tabular encoder.

#### 4.3.1 Tabular Encoder

TabPFN and its descendants [20, 36, 37] are a family of powerful tabular foundation models pretrained using synthetic data. TabPFN follows an in-context learning paradigm, where conditioning on a task-specific training set is required to obtain meaningful embeddings for multimodal learning, simultaneously enabling its predictive capacity. As a result, embedding extraction inherently produces the corresponding task predictions. Our use of TabPFN is therefore two-fold: First, we use TabPFN’s predictions as the clinical data-only output of our risk predictor. Formally, given a training set 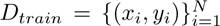, TabPFN generates *M* versions of *D_train_* by applying dataset permutation methods and feature transformations. The tabular risk estimation is thus defined as the mean over the *M* forward passes,

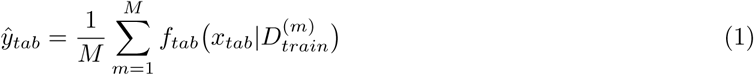

where *f_tab_* stands for the TabPFN model with its pretrained weights and *x_tab_* represents the input clinical data attributes.

Second, we use TabPFN to extract clinical data embeddings for multimodal fusion. Specifically, using the embedding extracting function *h*(*·, ·*) of the tabular encoder *f_tab_*, we extract the tabular representation *v_tab_* by concatenating the embeddings obtained via prompting the model using *M* version of the training data:

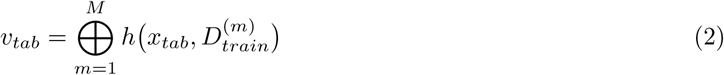

Concatenating embeddings across these perturbed versions yields stronger and more expressive clinical data representations, as described in [38].

#### 4.3.2 Multimodal Fusion

Incorporating TabPFN [20] as the tabular encoder allowed us to extend the predictive capabilities of our vision-tabular CVD risk model to tabular data-only scenarios. We follow recent vision-tabular architecture [18] to allow for strong CMR-only and CMR-tabular predictions, robust to missing values. Namely, we incorporate gated cross-attention as the modality fusion component for effective CMR-only and CMR-tabular predictions. The gated cross-attention module allows to adaptively adjust the weight of clinical data attributes depending on their amount and importance. CMR features *v_cmr_*are extracted from a vision encoder *f_cmr_*. Both CMR embeddings and tabular embeddings are then projected into a shared dimensional space *d*, resulting in *v*^*_cmr_* and *v*^*_tab_*. CMR embeddings are processed with self-attention, *v̂_cmr_* = *Self Attention*(*ṽ_cmr_*) and cross-attention is applied with image tokens *v̂_cmr_* as queries and tabular tokens *v*^*_tab_* as keys/values. A multilayer perceptron (MLP) is employed for the gate, which produces weights *w ∈* [0, 1] for the clinical data contribution, yielding a multimodal embedding, *v_multi_* = *v*^*_cmr_* + *w ⊙* CrossAttention(*v*^*_cmr_, v̂_tab_*). This representation is then passed through a feed-forward layer for downstream prediction.

#### 4.3.3 Two-Stage Training

To achieve optimal performance across all the possible inference scenarios, namely tabular-only, CMR-only, and CMR-tabular, we use a two-stage training procedure. As tabular-only performance is determined by the TabPFN model [20], which parameters remain frozen at all stages, our training focuses on optimizing CMR-only and multimodal predictions. The first stage of the training leaves all the trainable components of the method unfrozen. The model weights achieving the best CMR-only validation performance are then selected for the second stage. In this stage, only the cross-attention layers and gating mechanisms of the multimodal module remain trainable. This design preserves high CMR-only performance while enabling the multimodal components to learn optimal feature combinations. Additionally, we employ the TabMoFe loss [18] in both training stages, which constructs two nested subsets of the clinical attributes, one contained within the other, and ranks performance according to the number of available variables. This design encourages robust performance across varying levels of tabular data availability.

#### 4.3.4 Specialized Model Evaluation

TabPFN performs in-context learning at inference time, allowing it to be conditioned on a task-specific training set without gradient-based retraining [20, 36, 37]. We leverage this property to evaluate *specialized* models: for a given evaluation attribute subset (e.g., basic attributes only), we condition TabPFN on that same subset both for its predictions and for the embeddings passed to the multimodal fusion module. Notably, the CMR encoder and cross-attention fusion module, trained on the full attribute set, remain unmodified during this swap. Evaluating in this specialized setting allows us to compare the performance against the baselines without introducing attribute missingness as a confounding factor.

#### 4.3.5 Implementation Details

We use a 3D ResNet-50 [25] as the image encoder and TabPFN 2.5 [20] with its pretrained weights as the tabular encoder. The image encoder is initialized using pretrained weights from [18]. We use *M* = 8 for TabPFN, which results in a tabular embedding of size 8*192. The image embedding is of size 2048. The models are trained for 100 epochs with early stopping. We perform a learning rate sweep covering *{*10*^−^*^3^, 3 *×* 10*^−^*^3^, 10*^−^*^4^, 3 *×* 10*^−^*^4^, 10*^−^*^5^, 3 *×* 10*^−^*^5^*}* for our method and all the trainable baselines and report performance corresponding to the best learning rate. We use a batch size of 256. We use the Platt method [39] to calibrate the vision-tabular model on the validation set.

### 4.4 Evaluation

#### 4.4.1 Metrics

We evaluated model performance using the AUROC to assess discrimination between participants who developed CVD and those who did not, as it is a standard metric for unbalanced classification problems. AUROC distributions were obtained via stratified bootstrapping with 1,000 resamples of the test set, and are reported as the mean and 95% confidence interval across resamples. To assess calibration, we additionally report the Brier score (Supplementary Fig. A4). Pairwise comparisons of AUROC between our method and baseline models were assessed using DeLong’s test [40], which accounts for the correlation between paired predictions evaluated on the same test cohort. Individual AUROC values and their 95% confidence intervals were derived from the same DeLong covariance estimate used for significance testing, ensuring internal consistency between reported point estimates and hypothesis tests. For the per-attribute analysis (Fig. 1d), the *p*-values were corrected for multiple testing using the Benjamini-Hochberg false discovery rate procedure [41].

#### 4.4.2 Baselines

##### Traditional Risk Scores

The first group of baselines we use are established CVD risk scores, namely we compare against Framingham [3] and SCORE2 [4]. Those two risk scores have been derived using different geographical cohorts, Framingham being derived on a US population while SCORE2 on a European population. A list of attributes used for both the scores is available in Appendix Tabel A4. Both scores estimate 10-year risk of CVD, whereas our cohort’s outcome definition is based on the maximum observed time from imaging to incident CVD event in our cohort, which is shorter than 10 years (Section 4.2). To enable a comparable evaluation, we rescaled the baseline survival function of each risk score to match our outcome window, following the proportional hazards assumption *S*_0_(*t*) = *S*_0_(10)*^t/^*^10^, where *S*_0_(10) is the original 10-year baseline survival probability [24]. We use the published Framingham coefficients from [3] and the SCORE2 coefficients from [4], separated by sex as specified in the original derivations. As both scores were developed for specific age ranges (Framingham: 30-79 years; SCORE2: 40-69 years), we apply them to our full cohort without age restriction to ensure consistent comparison conditions across all evaluated methods. As Framingham and SCORE2 are not capable of handling missing values, we imputed missing attributes using the mode for categorical variables and the mean for numerical variables, computed from the training set.

##### Tabular Machine Learning Models

The second group of baselines are machine learning-based tabular models: Cox PH [24], XGBoost [42], and LightGBM [43]. As Cox PH is not capable of handling missing values, we imputed missing attributes in the same way as for traditional risk scores, using the mode for categorical variables and the mean for numerical variables, computed from the training set. LightGBM is implemented using the lightgbm python package, while XGBoost using the xgboost package. Both models are optimized using Optuna with 100 trials for each model. The details on the hyperparameter space are available in the Appendix Tabel A5. Cox PH was implemented using lifelines. When trained on broader clinical attribute sets, the Cox PH fitter encountered convergence failures; we therefore restrict its evaluation to the basic attribute set. As Cox PH produces a continuous risk score rather than a probability estimate, we use the predicted risk score directly to compute AUROC, treating higher scores as indicative of greater risk.

## Data availability

The UK Biobank data supporting the findings of this study are available-subject to a registration process and project approval on www.ukbiobank.ac.uk. This research has been conducted using the UK Biobank Resource under Application Number 87065.

## Code availability

The code will be made publicly available on GitHub upon the acceptance of the manuscript.

## Acknowledgments

M.H. is in part supported by the Munich School of Data Science (MUDS) and the European Laboratory for Learning and Intelligent Systems (ELLIS) PhD program. L.D. and M.H. are supported by the German Federal Ministry of Research, Technology and Space (DECIPHER-M, 01KD2420G).

## Author contributions

M.H. and J.A.S. designed the study. M.H. conceived the experiments, conducted the implementation and analysis, and wrote the manuscript. L.D. and M.D.F. contributed to the methodology and analysis of the results. M.H. and L.D. have access to all the data used in this study. K.B. provided medical interpretation of the results. J.A.S. provided supervision. All authors contributed to revising the manuscript and approved the final version.

## Appendix A Supplementary Materials

**Table A1.** List of attributes that achieved significant results against age-only baseline (AUROC 0.654 *±* 0.023) (Fig. 1D Main Paper). Values in parentheses indicate the AUROC improvement (*↑*) over the age-only baseline.

| Attribute | Category | AUROC | p-value |
| --- | --- | --- | --- |
| LV mean myocardial wall thickness AHA 10 | IDP | 0.709 ($\uparrow 0.055$) | 0.025 |
| LV mean myocardial wall thickness global | IDP | 0.710 ($\uparrow 0.056$) | 0.030 |
| LV mean myocardial wall thickness AHA 9 | IDP | 0.710 ($\uparrow 0.055$) | 0.017 |
| LV mean myocardial wall thickness AHA 1 | IDP | 0.707 ($\uparrow 0.052$) | 0.048 |
| Waist circumference | Physical Measures | 0.676 ($\uparrow 0.022$) | 0.048 |
| LV mean myocardial wall thickness AHA 7 | IDP | 0.710 ($\uparrow 0.055$) | 0.017 |
| LV circumferential strain AHA 13 | IDP | 0.678 ($\uparrow 0.024$) | 0.017 |
| Father’s age at death | Family History | 0.680 ($\uparrow 0.026$) | 0.030 |
| LVEDM | IDP | 0.708 ($\uparrow 0.054$) | 0.025 |
| LV mean myocardial wall thickness AHA 12 | IDP | 0.705 ($\uparrow 0.051$) | 0.048 |
| MYOESV | IDP | 0.709 ($\uparrow 0.055$) | 0.025 |
| MYOEDV | IDP | 0.707 ($\uparrow 0.053$) | 0.026 |

**Fig. A1.**
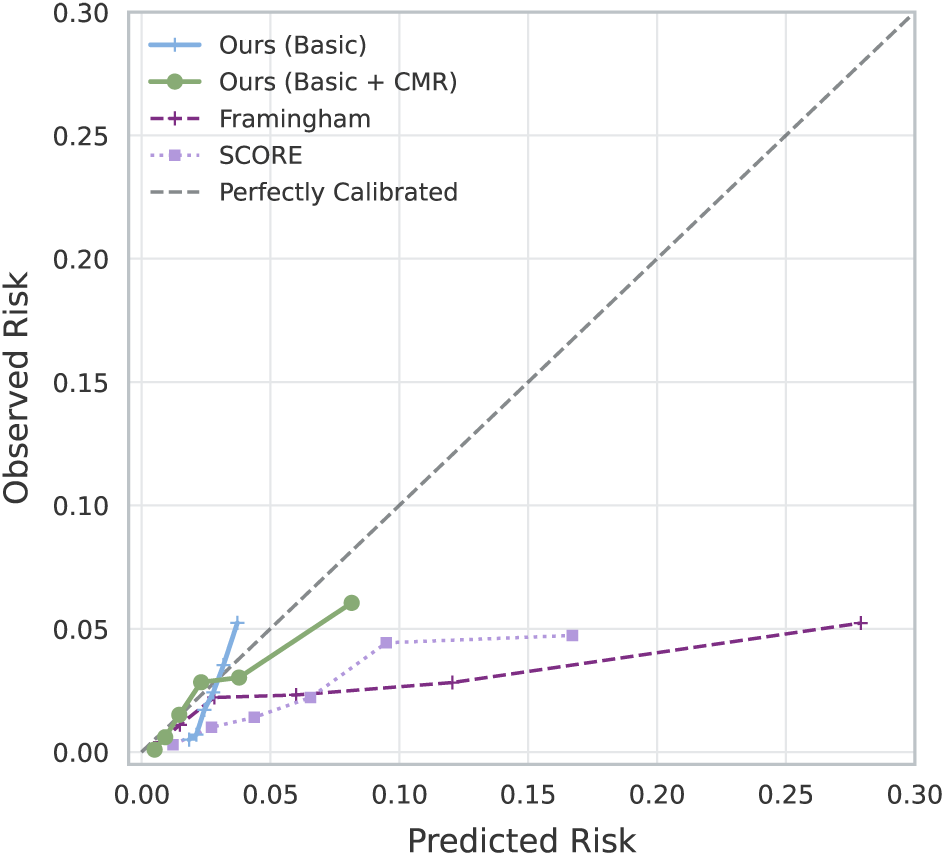
Calibration curves (extended). Predicted versus observed CVD risk for our vision-tabular model (Basic and Basic + Raw CMR variants), SCORE2, and Framingham, shown here with Framingham included for completeness (omitted from the main text for visual clarity).

**Table A2.** Attribute list per set. Complete list of clinical attributes included in the study, organized by attribute group. The listed groups collectively form the non-CMR attribute set. IDPs are listed separately in Appendix Tabel A3.

| Attribute Group | Attribute List |
| --- | --- |
| Basic CVD risk factors | Age, sex, total cholesterol, HDL cholesterol, systolic BP, BP medication, smoking status, diabetes |
| Family history | Father still alive, father's age at death, mother still alive, mother's age at death, non-accidental death in close genetic family, father has hip fracture, father has prostate cancer, father has severe depression, father has Parkinsons disease, father has Alzheimers disease/dementia, father has diabetes, father has high blood pressure, father has chronic bronchitis/emphysema, father has breast cancer, father has bowel cancer, father has lung cancer, father has stroke, father has heart disease, mother has hip fracture, mother has severe depression, mother has Parkinsons disease, mother has Alzheimers disease/dementia, mother has diabetes, mother has high blood pressure, mother has chronic bronchitis/emphysema, mother has breast cancer, mother has bowel cancer, mother has lung cancer, mother has stroke, mother has heart disease, siblings have hip fracture, siblings have prostate cancer, siblings have severe depression, siblings have Parkinsons disease, siblings have Alzheimers disease/dementia, siblings have diabetes, siblings have high blood pressure, siblings have chronic bronchitis/emphysema, siblings have breast cancer, siblings have bowel cancer, siblings have lung cancer, siblings have stroke, siblings have heart disease |
| Physical measurements | Height, weight, BMI, basal metabolic rate, body fat percentage, body surface area, diastolic blood pressure automated, hip circumference, diastolic blood pressure manual reading, waist circumference, whole body fat-free mass, whole body fat mass, whole body water mass, pulse rate automated, pulse wave arterial stiffness index |
| Lifestyle | Alcohol drinker status, alcohol intake frequency, current tobacco smoking, duration of heavy DIY, duration of strenuous sports, duration walking for pleasure, ever smoked, frequency of other exercises in last 4 weeks, days/week moderate activity, days/week vigorous activity, smoking/smokers in household, days/week walked, Duration of moderate activity, duration of vigorous activity, duration of walks, exposure to tobacco smoke at home, exposure to tobacco smoke outside home, pack years adult smoking as proportion of lifespan exposed, pack years of smoking, sleep duration |
| Medical history | Oral contraceptive pill or minipill, hormone replacement therapy, insulin, cholesterol lowering medication, overall health rating, pacemaker, shortness of breath walking on level ground, sleeplessness / insomnia, falls in the last year, worrier / anxious feelings, cardiac operations performed |
| PWA parameters | Augmentation index for PWA, cardiac index during PWA, cardiac output during PWA, central augmentation pressure during PWA, central pulse pressure during PWA, central systolic blood pressure during PWA, end systolic pressure during PWA, end systolic pressure index during PWA, diastolic brachial blood pressure during PWA, heart rate during PWA, mean arterial pressure during PWA, number of beats in waveform average for PWA, peripheral pulse pressure during PWA, stroke volume during PWA, systolic brachial blood pressure during PWA, total peripheral resistance during PWA |
| ECG conduction metrics | Ventricular rate, P duration, QRS duration |

**Table A3.**
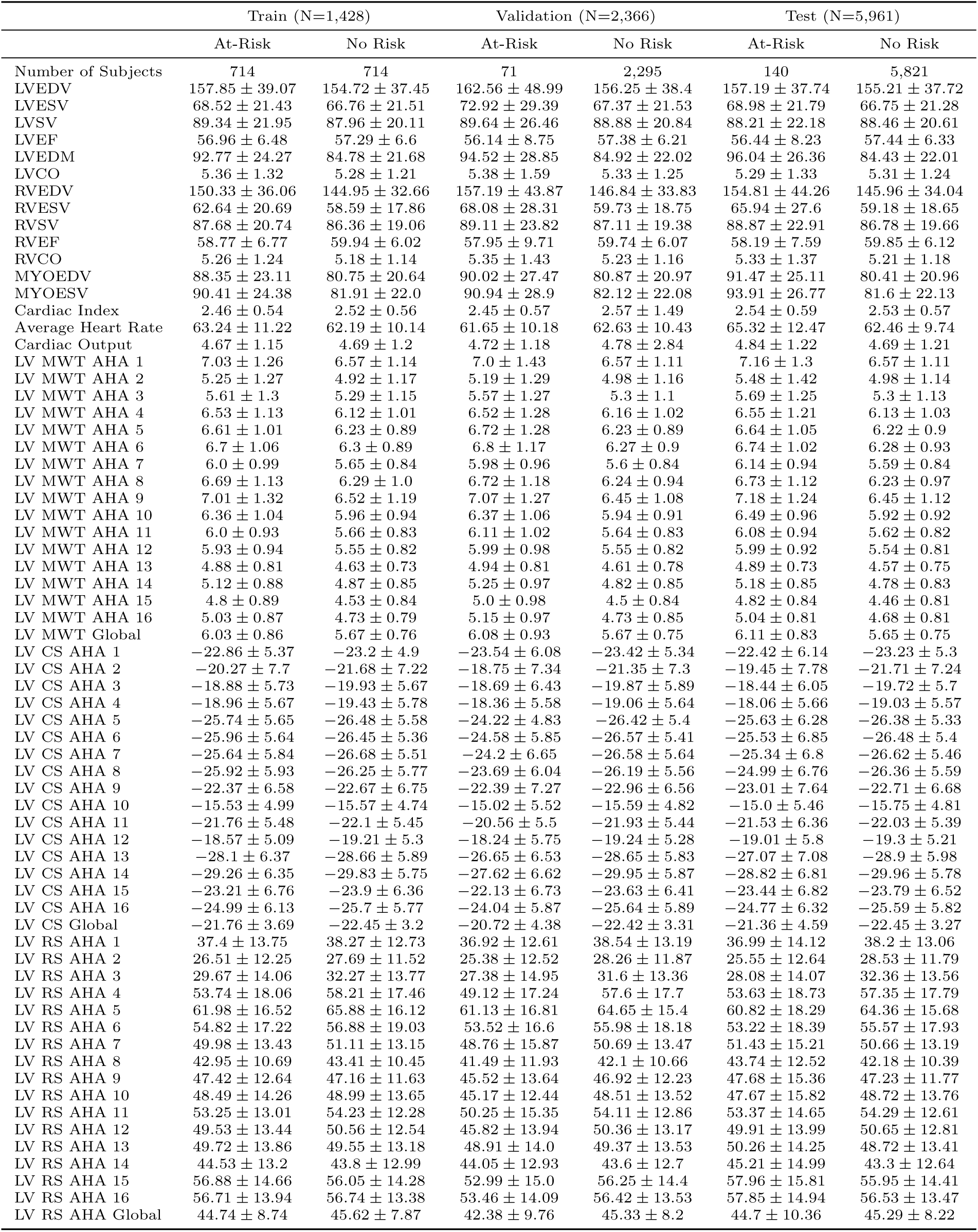
Image-derived phenotype (IDP) characteristics stratified by CVD risk status across dataset splits. Mean*±*standard deviation for all IDPs across training, validation, and test splits, separately for at-risk and no-risk participants. MWT: Myocardial Wall Thickness, CS: Circumferential Strain, RS: Radial Strain.

**Fig. A2.**
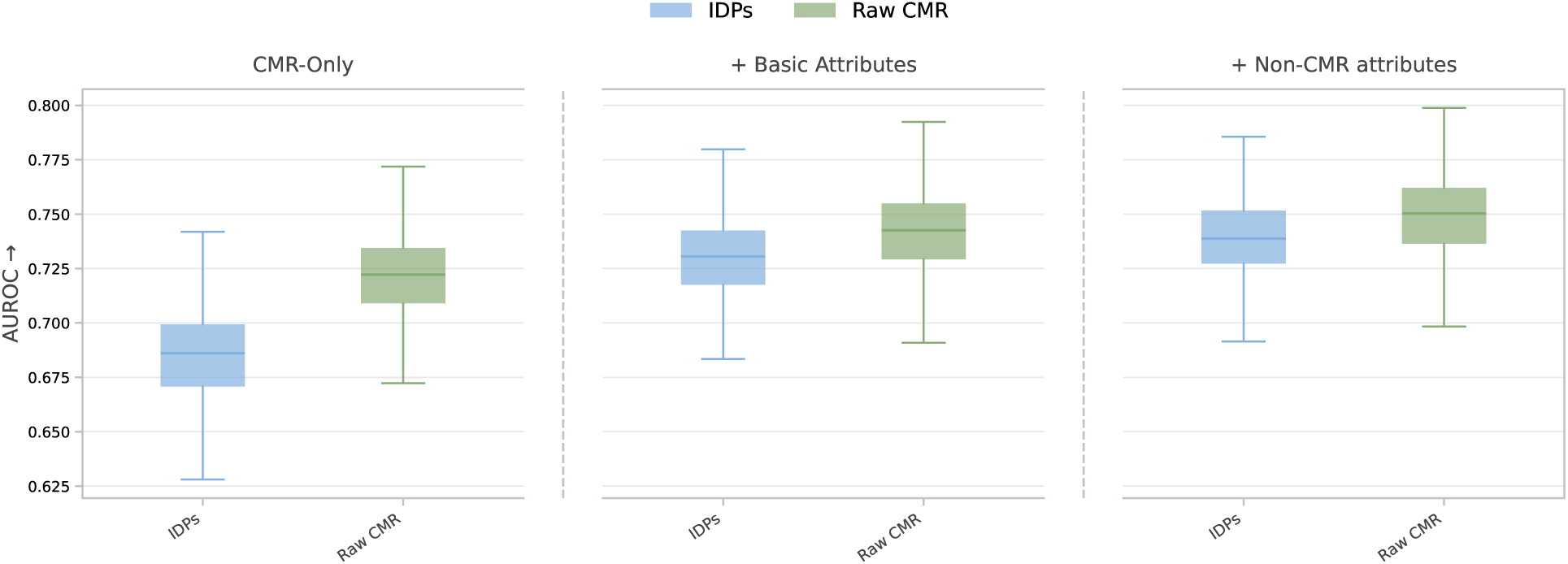
Comparison of predictive capability of CMR against IDPs. AUROC distributions across bootstrapped test set resamples comparing IDPs and raw cine CMR across three settings of increasing attribute availability: imaging only (CMR-Only), imaging combined with basic clinical attributes (+ Basic Attributes), and imaging combined with all non-CMR clinical attributes (+ Non-CMR Attributes). Raw cine CMR consistently matches or exceeds IDP performance across all settings, with the gap most pronounced in the imaging-only setting, suggesting that the end-to-end learned representations capture prognostic information beyond what is encoded in IDPs.

**Fig. A3.**
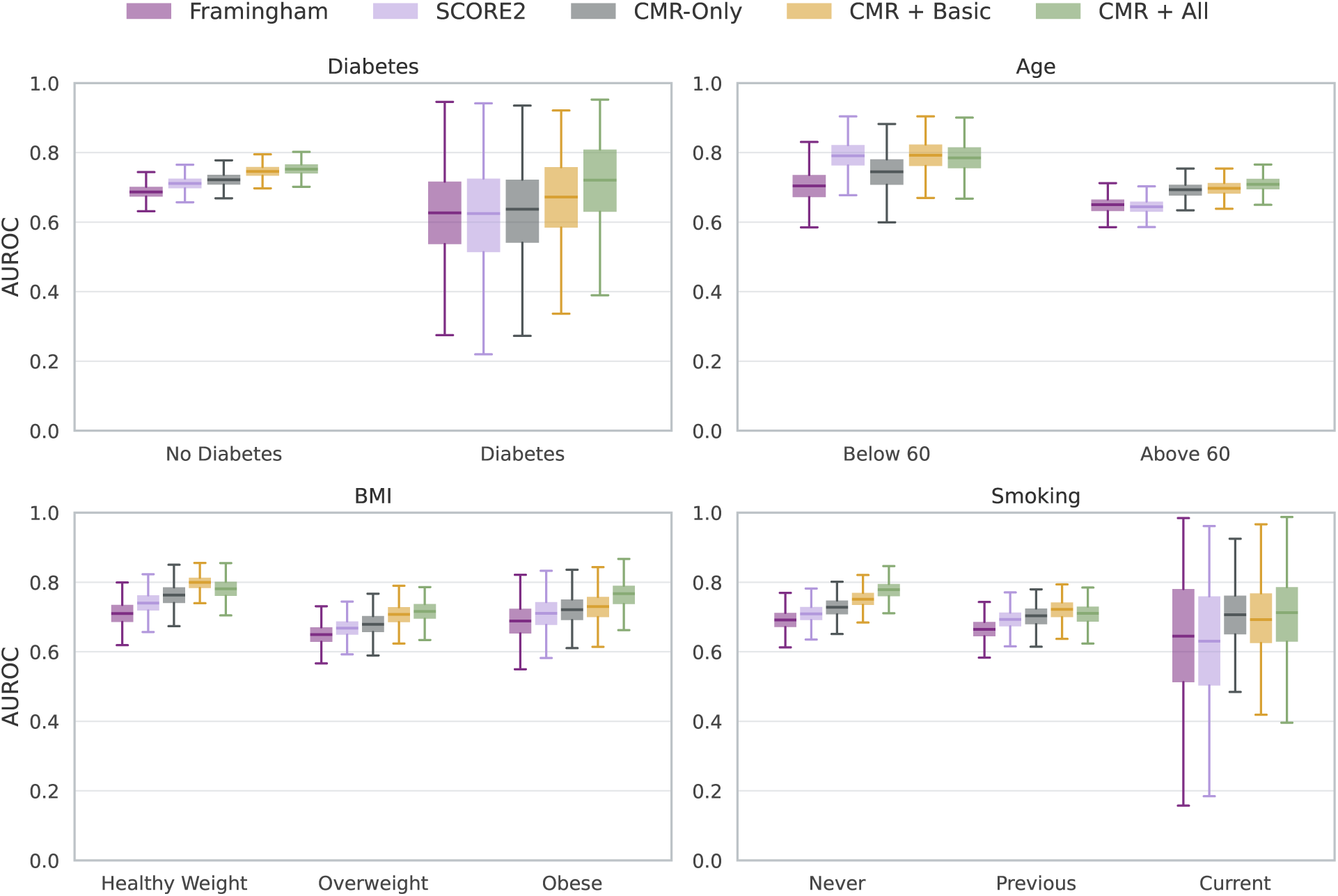
Subgroup analysis. AUROC distributions across bootstrapped test set resamples for Framingham, SCORE2, and CMR-based models (CMR-Only, CMR + Basic, CMR + All) stratified by diabetes (no diagnosed diabetes, diagnosed diabetes), age (below 60, above 60), BMI category (healthy weight, overweight, obese), and smoking status (never, previous, current). CMR-based models consistently match or outperform traditional risk scores across most subgroups. Performance is generally higher in women than men and in participants below 60. The current smoker subgroup shows substantially higher variance across all models, reflecting the small proportion of current smokers in the test set (approximately 3% of both at-risk and healthy participants), and results for this subgroup should therefore be interpreted with caution.

**Fig. A4.**
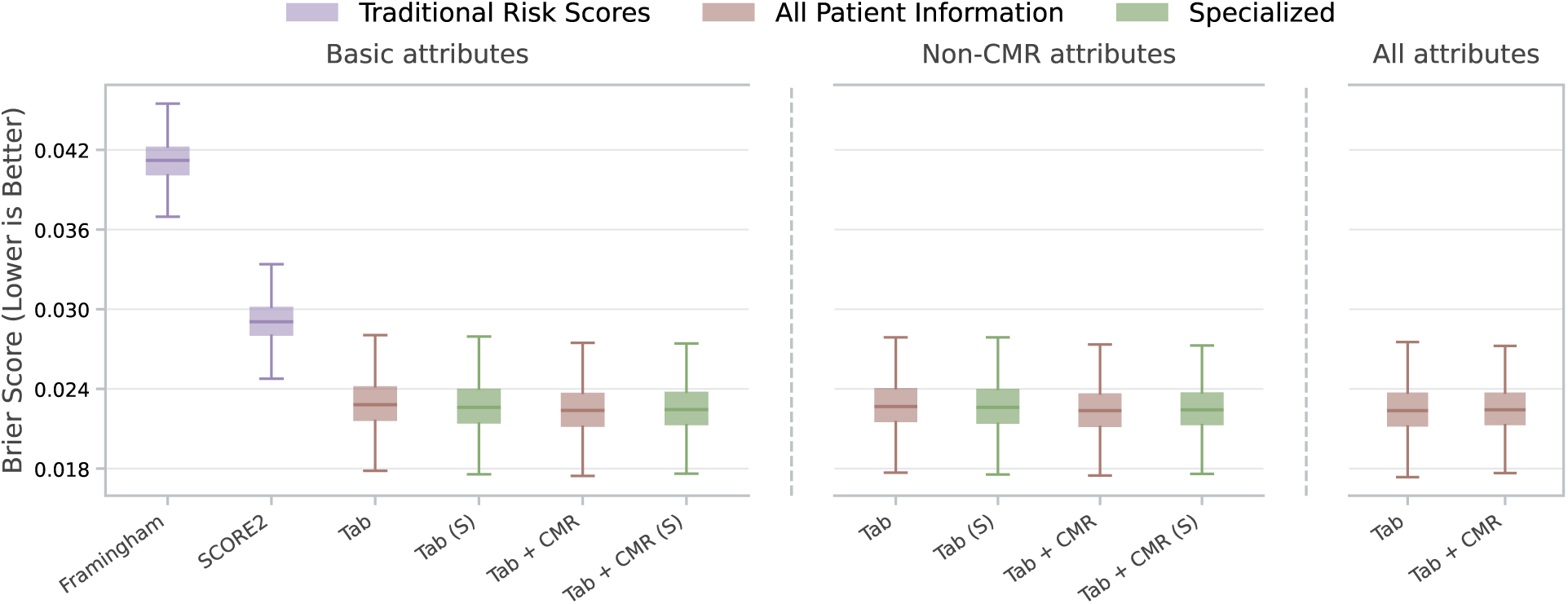
Calibration performance. Brier score distributions across bootstrapped test set resamples for all evaluated models, stratified by attribute set (basic, non-CMR, and all attributes). Lower values indicate better calibration. Traditional risk scores (Framingham, SCORE2) are shown only for the basic attribute set, as they operate on a fixed input schema. Tab and Tab + CMR refer to models trained on all available attributes and evaluated on the indicated subset, while Tab (S) and Tab + CMR (S) denote specialized models trained and evaluated on the same attribute subset. Across all attribute sets, Tab + CMR achieves competitive calibration with its tabular-only counterpart, while both substantially outperform traditional risk scores on the basic attribute set.

**Table A4.**
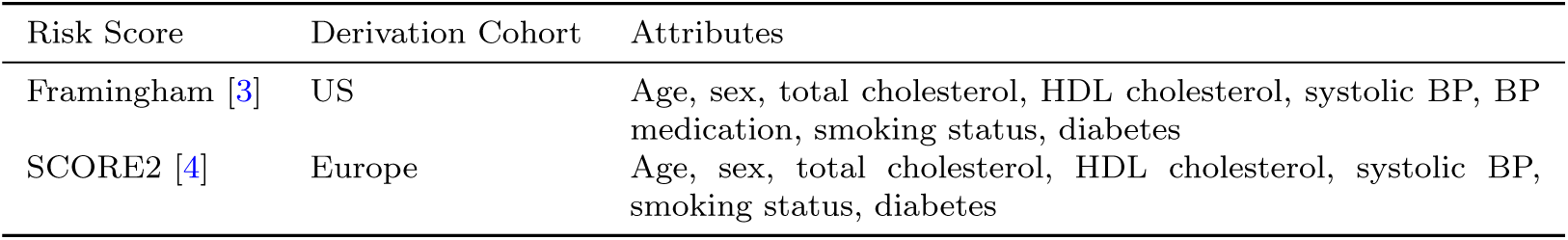
Traditional risk scores. Input attributes and derivation cohorts for the two established CVD risk scores used as baselines.

**Table A5.** Hyperparameter search space for tabular machine learning baselines (200 Optuna trials per model). Hyperparameter ranges and sampling distributions used during Optuna optimization for XGBoost [42] and LightGBM [43]. All searches were performed independently per model using the training set, with model selection based on validation AUROC.

| Model | Hyperparameter | Range | Distribution |
| --- | --- | --- | --- |
| XGBoost | max_depth | 2–10 | Integer, uniform |
|  | min_child_weight | 1–50 | Integer, uniform |
|  | gamma | 0.0–5.0 | Float, uniform |
|  | subsample | 0.7–0.9 | Float, uniform |
|  | colsample_bytree | 0.3–0.7 | Float, uniform |
|  | colsample_bylevel | 0.5–1.0 | Float, uniform |
|  | colsample_bynode | 0.5–1.0 | Float, uniform |
|  | eta (learning rate) | 0.01–0.1 | Float, log-uniform |
|  | lambda (L2 reg.) | 1.0–20.0 | Float, log-uniform |
|  | alpha (L1 reg.) | 0.0–10.0 | Float, uniform |
|  | max_bin | 128–512 | Integer, uniform |
| LightGBM | max_depth | 2–10 | Integer, uniform |
|  | min_data_in_leaf | 1–50 | Integer, uniform |
|  | min_split_gain | 0.0–5.0 | Float, uniform |
|  | bagging_fraction | 0.85–1.0 | Float, uniform |
|  | feature_fraction | 0.8–1.0 | Float, uniform |
|  | feature_fraction_bynode | 0.8–1.0 | Float, uniform |
|  | learning_rate | 0.01–0.1 | Float, log-uniform |
|  | reg_lambda (L2 reg.) | 1.0–20.0 | Float, log-uniform |
|  | reg_alpha (L1 reg.) | 0.0–10.0 | Float, uniform |

